# Alterations in Neuroimmune, Metabolic, and Oxidative (NIMETOX) Pathways in Post-Stroke Patients: Diagnostic and Prognostic Implications

**DOI:** 10.1101/2025.08.11.25333470

**Authors:** Thitiporn Supasitthumrong, Abbas F. Almulla, Chavit Tunvirachaisakul, Ana Paula Michelin, Andressa Keiko Matsumoto, Decio S. Barbosa, Elizabet Dzambazova, Yingqian Zhang, Michael Maes

## Abstract

**Background:** Stroke remains a primary contributor to disability and mortality globally. Survivors often experience persistent problems associated with neuro-immune, metabolic, and oxidative stress (NIMETOX) pathways. Nevertheless, the combined effects of these NIMETOX pathways on post-stroke functional outcomes have not been extensively investigated.

**Objectives:** The aim of this study is to examine alterations in NIMETOX pathways among stroke survivors relative to healthy controls. Additionally, this study aims to elucidate the predictive significance of NIMETOX biomarkers regarding stroke severity, disability, and functional outcomes.

**Methods:** A total of 40 healthy controls and 122 stroke survivors participated in the study. The NIH Stroke Scale (NIHSS), modified Rankin Scale (mRS), and Barthel Index (BI) were employed to evaluate stroke severity, disability, and functional independence, respectively.

**Results:** Findings indicate that immune-inflammatory markers, metabolic indices, and oxidative and nitrosative stress markers exhibited significant abnormalities in post-stroke patients. Binary logistic regression shows that increased atherogenicity, lipid hydroperoxides, and aberrations in the equilibrium between two catalytic sites of the paraoxonase 1 enzyme demonstrated high accuracy for stroke versus controls (93.9%). Regression analyses indicated that a combination of immune markers (increased interleukin-6, tumor necrosis factor-α, neutrophil-to-lymphocyte ratio), metabolic variables (Castelli risk index-1 and glycated hemoglobin), and increased lipid hydroperoxides significantly explained stroke severity (44.6% of the variance), disability (55.9%), and functional impairment (53.1%). Notably, IL-6 and TNF-α emerged as strong predictors of short-term stroke outcome.

**Conclusion:** Post-stroke functional deficits are largely predicted by the combined effects of immune and metabolic pathways, increased oxidative stress, and antioxidant disbalances. Treatment and prevention of stroke should target the different NIMETOX pathways.

## Introduction

Stroke represents a significant global health issue, especially in low- and middle-income countries, serving as a leading cause of mortality, disability, and economic burden (Katan & Luft, 2018). Although age-standardized stroke mortality has decreased, the global population of stroke survivors has risen, exceeding 25 million, with acute ischemic stroke (AIS) constituting most of these instances (Feigin *et al*., 2015; Meschia & Brott, 2018). Around 33% of these patients do not survive (Koton *et al*., 2010). Survivors frequently endure lasting physical disabilities, cognitive impairments, chronic fatigue, and mood disorders (Mitchell *et al*., 2017; Alghamdi *et al*., 2021; Weterings *et al*., 2023). Understanding the biological pathways that affect post-stroke outcomes is crucial for improving patient management and rehabilitation strategies.

Various studies highlight the critical roles of immune-inflammatory, metabolic, oxidative, and nitrosative stress pathways in influencing stroke progression and prognosis (Alfieri *et al*., 2020; Simats & Liesz, 2022). Following a stroke, microglial activation triggers the release of pro-inflammatory cytokines, notably tumor necrosis factor-alpha (TNF-α), interleukin (IL)-1β, and IL-6, detectable in brain tissue, cerebrospinal fluid, and peripheral circulation (Vila *et al*., 2000; Smith *et al*., 2004). Elevated IL-6 concentrations correlate with clinical deterioration, heightened stroke severity, greater disability, and unfavorable long-term outcomes (Whiteley *et al*., 2009; Shaafi *et al*., 2014). Elevated systemic inflammatory biomarkers, including high-sensitivity C-reactive protein (hsCRP), white blood cell (WBC) counts, and erythrocyte sedimentation rate (ESR), are predictive of negative post-stroke outcomes, such as disability and mortality (Furlan *et al*., 2014; Alfieri *et al*., 2020; Nguyen *et al*., 2020).

Oxidative and nitrosative stress play a crucial role in the pathophysiology following stroke. Post-stroke patients frequently exhibit abnormal levels of oxidative stress indicators, including reactive oxygen species (ROS), nitric oxide metabolites (NOx), lipid peroxidation products, and diminished antioxidant defenses (Manolescu *et al*., 2011; Maes *et al*., 2023). Redox disturbances adversely affect cerebral vasculature, increasing the likelihood of recurrent stroke and demonstrating a significant correlation with neurological severity (Ferretti *et al*., 2008; Maes *et al*., 2023). Reduced activity of antioxidant enzymes, especially paraoxonase-1 (PON1), is associated with stroke pathology, highlighting the therapeutic potential of antioxidant strategies aimed at oxidative stress and lipid peroxidation pathways (Ferretti *et al*., 2008; Brinholi *et al*., 2023). Brinholi et al. identified a significant correlation between AIS and PON1 activity indices, specifically CMPAase and AREase, suggesting their potential utility in diagnostics (Brinholi *et al*., 2023). Lipid hydroperoxide (LOOH) levels were significantly increased in both stroke survivor and non-survivor groups three months post-AIS when compared to control subjects (Reiche *et al*., 2019).

Metabolic dysregulation is a significant factor linked to the prognosis of IS. Elevated fasting blood glucose, glycated haemoglobin (HbA1c), triglycerides (TG), and total cholesterol (TC), coupled with reduced high-density lipoprotein cholesterol (HDL-C), are consistently associated with increased stroke severity, impaired functional recovery, and heightened mortality risk (Benjamin *et al*., 2018; Ali *et al*., 2024). Lower HDL-C levels at the time of stroke admission are significantly associated with greater clinical severity and worse functional outcomes (Yeh *et al*., 2013).

Maes et al.(2025) recently defined the term “NIMETOX,” synthesizing various findings that highlight the interrelated abnormalities in neuroimmune, metabolic, and oxidative stress pathways associated with major depression (Maes *et al*., 2025a; Maes *et al*., 2025b). Numerous studies have explored individual abnormalities in immune, metabolic, and oxidative stress pathways in stroke populations (Kawabori & Yenari, 2015; Lehmann *et al*., 2022; Brinholi *et al*., 2023; Maes *et al*., 2023). However, a comprehensive examination of the NIMETOX pathways, namely metabolic and oxidative stress pathways with cytokine networks, in post-stroke patients is currently lacking. Understanding whether cytokine networks play a role in post-stroke disabilities above and beyond the effects of metabolic and oxidative stress pathways could inform personalized therapeutic strategies to enhance recovery and minimize stroke recurrence.

Thus, the objective of this study is to examine whether cytokine networks impact post-stroke disabilities above and beyond the effects of other NIMETOX biomarkers (Maes *et al*., 2023) including immune (hsCRP, WBC, neutrophil-to-lymphocyte ratio), metabolic (HDL-C, TC, TG, fasting blood glucose, HbA1c), and oxidative/nitrosative stress (PON1 enzyme activities [CMPAase, AREase], thiol groups, nitric oxide metabolites [NOx], and lipid hydroperoxides) biomarkers. Therefore, this study examines the cumulative effects of the NIMETOX pathways on clinical outcomes, such as short-term stroke severity (assessed by the National Institutes of Health Stroke Scale; NIHSS), overall disability (measured using the modified Rankin Scale; mRS), and functional independence (evaluated with the Barthel Index). We hypothesize that post-stroke patients would exhibit significant aberrations in cytokine networks that coupled to metabolic and oxidative stress pathways strongly correlate with stroke severity and functional outcomes.

## Method

### Participants

All participants gave informed consent, and the study was approved by the Institutional Review Board of the Faculty of Medicine, Chulalongkorn University, in accordance with the Declaration of Helsinki and ICH-GCP standards (IRB no. 62/073).

We employed a case-control design to investigate the biological underpinnings of post-stroke functional deficits by examining inflammatory cytokines, metabolic variables, and oxidative stress biomarkers. The oxidative stress biomarkers and metabolic markers have been published before (Maes *et al*., 2023), although in the present study we introduce more comprehensive metabolic indices reflecting Castelli risk index 1 and atherogenic index of plasma (AIP). Moreover, in part of the participants we assayed 27 cytokines, chemokines and growth factors to examine the effects of these immune markers in combination with metabolic and oxidative stress biomarkers on stroke outcome. Participants were recruited from the Stroke Unit at King Chulalongkorn Memorial Hospital during the period spanning from October 2019 to September 2020, specifically involving admitted patients. We included 122 patients diagnosed with AIS, based on focal neurological deficits persisting beyond 24 hours and verified through clinical examination and computed tomography imaging. Stroke subtypes were diagnosed according to the TOAST (Trial of ORG 10172 in Acute Stroke Treatment) criteria. In addition, 40 healthy control subjects, matched for age, sex, and educational background, were recruited from the general population in the same catchment area.

The criteria for patient exclusion were delineated as follows: (a) individuals who had experienced a transient ischemic attack or a hemorrhagic stroke, and (b) individuals who were unable to engage in the verbal clinical interview because they were unconscious or aphasic or suffering from a delirium. The criteria for exclusion of both patients and controls were delineated as follows: (a) cancer, liver failure, renal disease, infectious diseases, such as hepatitis B and HIV; (b) neurodegenerative or neuroinflammatory disorders such as multiple sclerosis, Parkinson’s disease, and Alzheimer’s disease, (c) immune disorders including inflammatory bowel disease, systemic lupus erythematosus, type 1 diabetes mellitus, chronic obstructive pulmonary disease, psoriasis, and rheumatoid arthritis, (d) lifetime psychiatric disorders including bipolar disorder, schizophrenia, major depressive disorder, substance use disorders, psycho-organic syndrome obsessive-compulsive disorder, and post-traumatic stress disorder, (e) use of psychoactive medications including antidepressants and antipsychotics.

### Clinical assessments

The clinical assessments encompassed the National Institutes of Health Stroke Scale (NIHSS), the Modified Rankin Scale (mRS), and the Barthel Index. The clinical assessments were performed within a 24-hour period following admission. The NIHSS serves as a critical tool for assessing the severity of a stroke, systematically evaluating key domains including language capabilities, dysarthria, levels of consciousness, facial paralysis, sensory perception, attentional deficits, motor drift in the arms and legs, limb ataxia, visual field integrity, and extraocular movements. mRS is commonly utilized in clinical settings to assess the extent of disability, overall impairment, or dependence of individuals. The Barthel index serves as a quantitative assessment of a patient’s autonomy in performing essential daily activities, including mobility, feeding, and bathing. In the present investigation, we employ two z unit-based composite scores designed to evaluate the severity of short-term stroke outcomes, specifically (a) z NIHSS + z mRS, and (b) z NIHSS + z mRS – z Barthel index within 48 hrs after stroke (Maes *et al*., 2023).

### Assays

Blood samples for the assessment of the NIMETOX indicators, were collected at 8:00 a.m. after an overnight fast and within 48 hours of hospital admission. Samples of serum were stored at a temperature of -80 °C until they were thawed for subsequent analysis. The metabolic biomarkers included total cholesterol, HDL-cholesterol, triglycerides, fasting blood glucose (FBG) and HbA1c. Employing the Alinity C (Abbott Laboratories, USA; Otawara-Shi, Tochigi-Ken, Japan), measurements of total cholesterol, HDL cholesterol, and triglycerides were conducted utilizing enzymatic techniques for total cholesterol, accelerator selective detergent methods for HDL cholesterol, and glycerol phosphate oxidase for triglycerides. The coefficients of variation observed for total cholesterol, HDL cholesterol, and triglycerides were recorded at 2.3%, 2.6%, and 2.3%, respectively. The inter-assay coefficients of variation for fasting blood glucose (FBG) and hemoglobin A1c (HbA1c) measured using the Alinity C (Abbott Laboratories, USA; Otawara-Shi, Tochigi-Ken, Japan) through an enzymatic technique were found to be 1.7% and 1.5%, respectively.

The biomarkers associated with oxidative stress that were measured encompass LOOH, NOx, -SH groups, chloromethyl phenylacetate (CMPA)ase, and arylesterase (AREase). NOx were quantified through the measurement of nitrite and nitrate concentrations utilizing a microplate reader (EnSpire®, Perkin Elmer, USA) at a wavelength of 540 nm. The -SH groups were measured using a microplate reader (En-Spire®, Perkin Elmer, USA) at a wavelength of 412 nm (Hu, 1994; Taylan & Resmi, 2010). The measurement of LOOH was performed using chemiluminescence with a Glomax Luminometer (TD 20/20). The procedure was conducted in a dark setting at a regulated temperature of 30 °C for a period of 60 minutes. The results were expressed as relative light units. The assessment of PON1 status was conducted via the activity of AREase and CMPAase, alongside the PON1 Q192R genotypes (Maes *et al*., 2023). An investigation was conducted on the rate of phenylacetate hydrolysis at low salt concentrations by evaluating the activity of AREase and CMPAase (Sigma, St. Louis, MO, USA)-ase. The experimental procedure employed a Perkin Elmer® EnSpire model microplate reader from Waltham, MA, USA, to monitor the hydrolysis rate of phenylacetate systematically. The experiment was performed at a controlled temperature of 25°C for a duration of 4 minutes, during which 16 measurements were recorded at 15-second intervals. The activity was measured in units per millilitre (U/mL), using the phenyl acetate molar extinction coefficient of 1.31 mMol/Lcm-1 as the measurement standard. The inter-assay coefficients of variation for the oxidative stress variables were all below 10.0%.

The immunological biomarkers evaluated comprised white blood cell (WBC) count, the neutrophil/lymphocyte ratio (NLR), and hsCRP. The assessment of these biomarkers was performed using the Alinity C system from Abbott Laboratories, located in Chicago, Illinois, USA, and Otawara-Shi, Tochigi-Ken, Japan. This study utilized a flow cytometric technique with a semiconductor laser to evaluate white blood cell count and an immunoturbidimetric method for the quantification of hsCRP. The laboratory has determined the following inter-assay coefficients of variation: hsCRP at 2.03%, WBC count at 2.0%, neutrophil percentage at 1.9%, and lymphocyte percentage at 3.5%.

The concentrations of cytokines, chemokines, and growth factors in plasma samples were determined using the Bio-Plex Pro human cytokine 27-plex test kit (BioRad, Carlsbad, California, USA). The LUMINEX 200 apparatus was utilized to measure the fluorescence intensities (FI) of the detecting antibodies and streptavidin-PE (BioRad, Carlsbad, California, USA). The intra-assay coefficient of variance for all cytokines was found to be below 11%. The Electronic Supplementary File (ESF), Table 1, enumerates the names, acronyms, and official gene symbols corresponding to all the cytokines, chemokines, and growth factors assessed in this study. IL-2, IL-5, IL-10, IL-15, G-CSF, GM-CSF and VEGF demonstrated a significant prevalence of results falling below the assay’s sensitivity threshold (exceeding 70%). Consequently, these cytokines were excluded from the analyses. IL-7, IL-12p70 and IL-17A showed measurable levels in more than 30% but less than 65% of the participants. Therefore, these three cytokines were entered into the analyses as prevalences (dummy variables). Consequently, we performed principal component analysis to extract significant networks (principal component) from the measurable cytokines (Maes *et al*., 2024). Cytokines, chemokines, and growth factors were employed, where needed, following fractional rank-based, logarithmic (log10), and square root transformations, or Winsorization, with all data subsequently processed as z transformations.

**Table 1.** Socio-demographic and clinical data in post-stroke patients and healthy controls.

| Variables | Healthy Controls<br>(n=40) | Post-Stroke<br>(n=122) | F / $\chi^2$ | df | p-value |
| --- | --- | --- | --- | --- | --- |
| Age (years) | 60.6 (9.2) | 60.1 (9.1) | 0.11 | 1/160 | 0.744 |
| Body mass index (kg/m<sup>2</sup>) | 24.90 (3.93) | 24.93 (3.82) | 0.00 | 1/160 | 0.963 |
| Gender (Male/Female) | 18/22 | 68/54 | 1.40 | 1 | 0.238 |
| Education level | 4.58 (1.97) | 4.66 (1.97) | 0.05 | 1/160 | 0.823 |
| FNDS score | 40/0 | 117/5 | 1.69 | 1 | 0.334 |
| Hypertension (No/Yes) | 40/0 | 40/82 | 54.44 | 1 | <0.001 |
| Diabetes mellitus (No/Yes) | 40/0 | 82/40 | 17.42 | 1 | <0.001 |
| Dyslipidemia (No/Yes) | 40/0 | 82/40 | 17.42 | 1 | <0.001 |
| Ischemic Heart Disease (No/Yes) | 40/0 | 114/8 | 2.76 | 1 | 0.097 |
| Stroke previous (No/Yes) | 40/0 | 99/23 | 8.79 | 1 | 0.003 |
| NIHSS (basal) | 0 (0) | 3.05 (1.88) | MWUT | - | <0.001 |
| mRS (basal) | 0 (0) | 2.18 (0.83) | MWUT | - | <0.001 |
| Barthel Index | 100 (0) | 85.56 (14.44) | MWUT | - | <0.001 |
| z NIHSS + z mRS (z score) | -1.28 (0) | 0.422 (0.776) | MWUT | - | <0.001 |
| z NIHSS + z mRS + z Barthel (z score) | -1.28 (0) | 0.425 (0.775) | MWUT | - | <0.001 |

FNDS: Fagerstrom Nicotine Dependence Scale; NIHSS: National Institutes of Health Stroke Scale; Barthel Index for Activities of Daily Living; mRS: Modified Rankin Score; MWUT: Mann-Whitney U test

#### Statistical analysis

Data analyses were performed utilizing IBM SPSS Statistics for Windows, Version 30. Differences between groups in continuous variables were assessed through analysis of variance (ANOVA), and relationships between categorical variables were analyzed using chi-square (χ²) tests. For the comparison of NIHSS, mRS, Barthel Index, z NIHSS + z mRS, and z NIHSS + z mRS + z Barthel, a Mann-Whitney U test (MWUT) was used. To mitigate the risk of Type I errors arising from multiple comparisons, p-values were adjusted utilizing the False Discovery Rate (FDR) correction. Pearson’s product-moment correlation coefficients were calculated to investigate relationships among continuous variables. Analyses were modified to account for essential demographic variables, such as age, sex, body mass index (BMI), and smoking status. Manual entry and stepwise regression procedures were utilized, with entry and removal thresholds established at p = 0.05 and p = 0.06, respectively. Statistics reported comprised standardized beta coefficients, degrees of freedom (df), p-values, R², and F-statistics. The assumptions of heteroskedasticity were evaluated through the application of the White test and a modified Breusch–Pagan test. Multicollinearity was assessed using tolerance values and variance inflation factors (VIFs). All tests were conducted as two-tailed, with statistical significance established at p < 0.05. Binary logistic regression was utilized to investigate the association between NIMETOX biomarkers and the probability of post-stroke diagnosis. In this model, post-stroke status was designated as the dependent variable, using healthy controls as the reference group. The analysis was adjusted for potential confounders such as age, sex, BMI, and smoking status. Effect sizes were calculated employing Nagelkerke’s pseudo-R². Outputs comprised Wald statistics with corresponding p-values, odds ratios (ORs) along with their 95% confidence intervals (CIs), and unstandardized regression coefficients (B) accompanied by standard errors (SE).

Principal component analysis (PCA) followed by varimax rotation was utilized to extract latent constructs from the cytokine dataset. The Kaiser–Meyer–Olkin (KMO) measure was utilized to evaluate sampling adequacy, with values exceeding 0.70 signifying adequate factorability. Principal components were retained if they accounted for over 50% of the variance and exhibited factor loadings exceeding 0.60 on all contributing variables.

## Results

### Sociodemographic and Clinical Features of Study Groups

**Table 1** demonstrates a comparison of the sociodemographic and clinical characteristics between post-stroke patients and healthy controls, including indices of disability and clinical severity such as NIHSS, mRS, and the Barthel Index scores. Patients who experienced a stroke exhibited significantly higher prevalence of comorbid conditions, notably diabetes mellitus (DM), hypertension (HT), and dyslipidemia (DysL), compared to the control group. Moreover, marked differences emerged between groups in terms of stroke history and scores on baseline clinical severity scales. Specifically, post-stroke patients displayed substantially elevated basal NIHSS and mRS scores, alongside reduced Barthel Index scores. Furthermore, the composite scores z NIHSS + z mRS and z NIHSS + z mRS – z Barthel were significantly elevated among stroke survivors. In contrast, the two groups showed no significant differences regarding age, BMI, gender distribution, educational attainment, tobacco use scores, or incidence of ischemic heart disease (IHD).

### Results of Factor Analyses

PCA was employed to discern potential underlying dimensions among the measurable proinflammatory cytokines: IL-1Ra, IL-4, IL-8, IL-9, Eotaxin, IP-10, MCP-1, MIP-1α, PDGF-BB, MIP-1β, RANTES, IL-1β, IL-6, IL-13, FGF, and TNF-α. The last 5 cytokines/growth factors did not load highly on the PCs extracted (all loadings < 0.45) and were, therefore, not included. **Table 2** presents the varimax-rotated first two PCs. These two PCs explained together 67.23% of the variance, with the first varimax-rotated PC explaining 38.65% of the variance and the second 28.58%. IL-1Ra, IL-4, Eotaxin, IP-10, MCP-1, PDGF-BB and RANTES loaded highly on the first PC (denoted as PC1). The second varimax-rotated PC loaded highly on IL-8, IL-9, MIP-1α and MIP-1β (denoted as PC2). As a result, we used PC1 score, PC2 score, IL-1β, IL-6, IL-13, FGF, and TNF-α, and the prevalences of IL-7, IL-12p70 and IL-17A (entered as dummy variables) in the analyses.

**Table 2.**
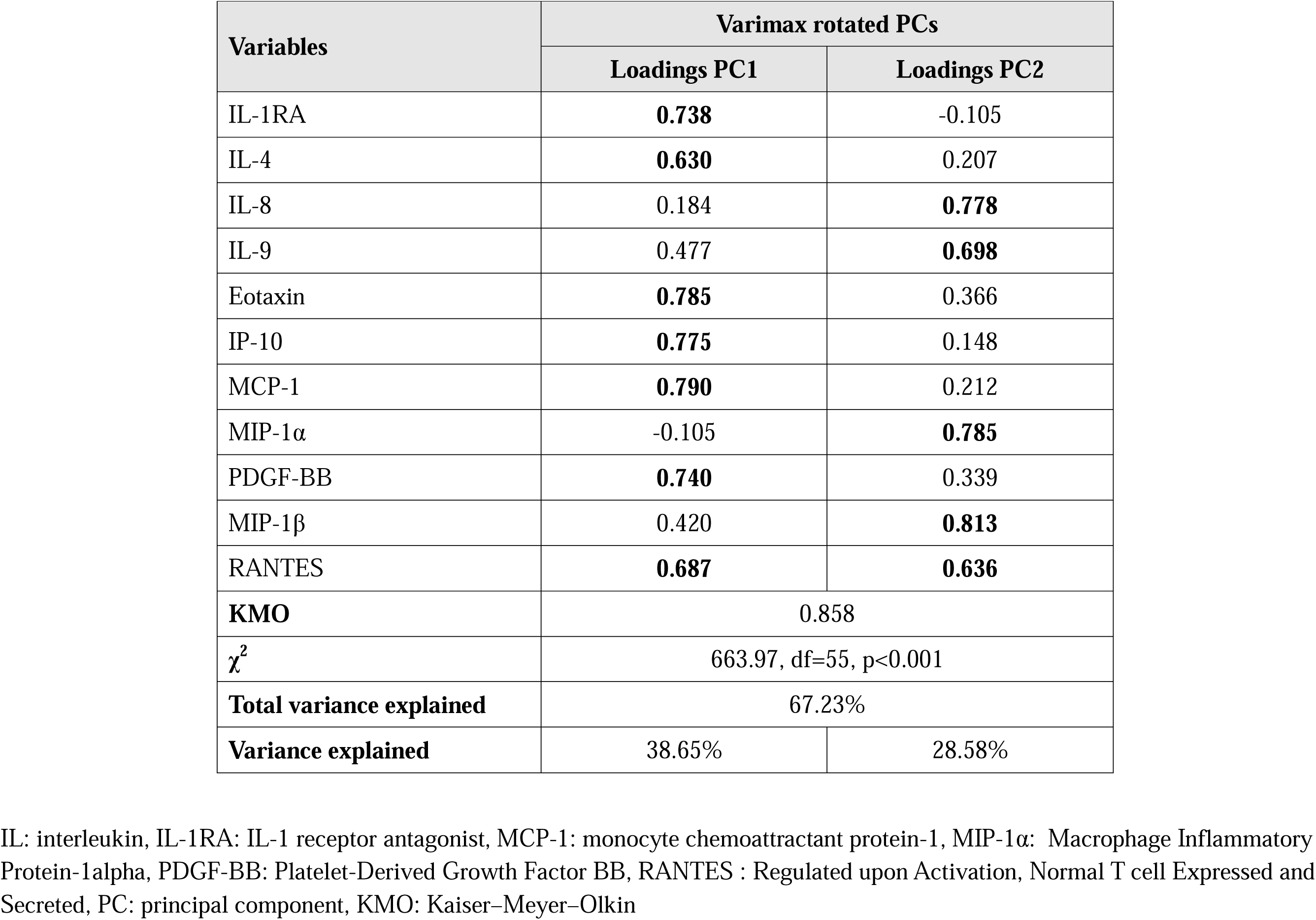
Results of principal component analysis (PCA) conducted on serum cytokine measurements.

| Variables | Varimax rotated PCs |  |
| --- | --- | --- |
|  | Loadings PC1 | Loadings PC2 |
| IL-1RA | **0.738** | -0.105 |
| IL-4 | **0.630** | 0.207 |
| IL-8 | 0.184 | **0.778** |
| IL-9 | 0.477 | **0.698** |
| Eotaxin | **0.785** | 0.366 |
| IP-10 | **0.775** | 0.148 |
| MCP-1 | **0.790** | 0.212 |
| MIP-1α | -0.105 | **0.785** |
| PDGF-BB | **0.740** | 0.339 |
| MIP-1β | 0.420 | **0.813** |
| RANTES | **0.687** | **0.636** |
| **KMO** | 0.858 |  |
| $\chi^2$ | 663.97, df=55, p<0.001 | |
| **Total variance explained** | 67.23% |  |
| **Variance explained** | 38.65% | 28.58% |

IL: interleukin, IL-1RA: IL-1 receptor antagonist, MCP-1: monocyte chemoattractant protein-1, MIP-1α : Macrophage Inflammatory Protein-1alpha, PDGF-BB: Platelet-Derived Growth Factor BB, RANTES : Regulated upon Activation, Normal T cell Expressed and Secreted, PC: principal component, KMO: Kaiser–Meyer–Olkin

### NIMETOX biomarkers in post-stroke patients compared to healthy controls

**Table 3** summarizes that post-stroke patients showed significant increases in various blood cell biomarkers, including total WBC count and NLR, when compared to healthy control participants. Post-stroke patients exhibited notable dysregulation in lipid metabolism, evidenced by significantly reduced HDL levels, as well as composite scores reflecting increased total cholesterol-to-HDL (TC/HDL) and triglyceride-to-HDL (TG/HDL) ratios.

**Table 3.** Results of general linear models (GLM) show the associations between biomarkers and post-stroke patients versus healthy controls (HC). The associations were adjusted for age, sex, BMI, and smoking.

| Variables | Healthy Controls (n=40) | Post-Stroke Patients (n=122) | F | df | p-value |
| --- | --- | --- | --- | --- | --- |
| High sensitivity C-reactive protein (mg/L) | 1.52(1.63) | 5.21(0.93) | 3.84 | 1/156 | 0.052 |
| White blood cell count (K/μL) | 5.50(0.35) | 8.21(0.20) | 44.66 | 1/156 | <0.001 |
| N/L ratio (NLR) | 1.64(0.33) | 2.71(0.19) | 7.80 | 1/156 | 0.006 |
| High-density lipoprotein (HDL) (mg/dL) | 58.8(1.9) | 45.4(1.1) | 38.34 | 1/156 | <0.001 |
| Total cholesterol (TC) (mg/dL) | 187.1(8.1) | 192.4(4.6) | 0.33 | 1/156 | 0.568 |
| Triglycerides (TG) (mg/dL) | 113.1(13.6) | 139.3(7.8) | 2.79 | 1/156 | 0.097 |
| z TC – z HDL ratio (z score) | -0.627(0.147) | 0.206(0.084) | 24.18 | 1/156 | <0.001 |
| z TG – z HDL ratio (z score) | -0.614(0.147) | 0.201(0.084) | 23.07 | 1/156 | <0.001 |
| Fasting Blood Glucose (mg/dL) | 108.4(8.0) | 129.6(4.6) | 5.22 | 1/156 | 0.024 |
| Glycated hemoglobin (%) | 5.94(0.30) | 6.99(0.17) | 9.49 | 1/156 | 0.002 |
| z CMPAase - z AREase (z score) | 0.849(0.136) | -0.288(0.079) | 32.04 | 1/152 | <0.001 |
| z CMPAase + z HDL (z score) | 0.618(0.147) | -0.218(0.085) | 23.99 | 1/152 | <0.001 |
| -SH groups (μmol/L) | 298.730(9.799) | 252.107(5.683) | 16.84 | 1/152 | <0.001 |
| Nitric oxide metabolites (μmol/L) | 7.405(0.584) | 5.467(0.339) | 8.19 | 1/152 | 0.005 |
| Log Lipid hydroperoxides (LOOH) (RLU) | 4.15(0.03) | 4.37(0.02) | 39.59 | 1/152 | <0.001 |
| PC1-cytokine score | -0.665(0.208) | 0.229(0.121) | 13.71 | 1/76 | <0.001 |
| PC2-cytokine score | 0.851(0.193) | -0.293(0.112) | 26.14 | 1/76 | <0.001 |
| Interleukin (IL)-1β (pg/mL) | 2.33(0.41) | 1.12(0.24) | 2.99 | 1/76 | 0.088 |
| IL-6 (pg/mL) | 2.48(2.52) | 4.89(1.47) | 2.52 | 1/76 | 0.117 |
| IL-13 (pg/mL) | 4.35(0.62) | 3.84(0.36) | 0.77 | 1/76 | 0.384 |
| Fibroblast Growth Factor-basic (pg/mL) | 5.26(0.67) | 2.64(0.40) | 10.09 | 1/76 | 0.002 |
| Tumor Necrosis Factor-α (pg/mL) | 5.52(1.00) | 4.88(0.59) | 0.29 | 1/76 | 0.592 |
| Prevalence IL-7 (no/yes) | 12/9 | 30/31 | 0.40 | 1 | 0.529 |
| Prevalence IL-12p70 (no/yes) | 10/11 | 23/38 | 0.64 | 1 | 0.424 |
| Prevalence IL-17A (no/yes) | 6/15 | 25/36 | 1.02 | 1 | 0.312 |

All results are shown as marginal estimated mean (SE); all results of GLM analyses with age, sex, BMI and smoking as covariates. CMPAase: 4-chloromethylphenylacetate, AREase: Arylesterase, PC: principal component

In terms of glycemic indices, HbA1c and fasting blood glucose levels were significantly elevated in post-stroke patients compared to controls. Post-stroke patients exhibited elevated levels of LOOH. In contrast, z CMPAase – z AREase, thiol groups, and NOx were significantly lower compared to healthy subjects.

Additionally, the PC1 score was significantly increased in the post-stroke group as compared to controls. The PC2 score and fibroblast growth factor-basic (FGF-basic) exhibited significantly reduced levels in post-stroke patients relative to healthy controls. No statistically significant differences were observed between the two groups for several cytokines, including IL-1β, IL-6, IL-13, and TNF-α, as well as for dummy-coded cytokine indicators such as IL-7, IL-12, and IL-17. Finally, no notable differences were detected between post-stroke patients and control participants regarding hsCRP, total cholesterol (TC), and triglycerides (TG). All the significant differences described above remained significant after FDR p-correction.

### Correlation Analysis

**Table 4** displays the correlation matrix that shows the relationships between different NIMETOX biomarkers and clinical measures of stroke severity. The NIHSS and mRS scores exhibited a positive and significant correlation with inflammation and oxidative stress markers, such as hsCRP, WBC count, NLR, TC/HDL and TG/HDL ratios, PC1, IL-6, and lipid hydroperoxides. In contrast, these severity indices exhibited an inverse relationship with HDL cholesterol, PC2s, z CMPAase – z AREase.

**Table 4.**
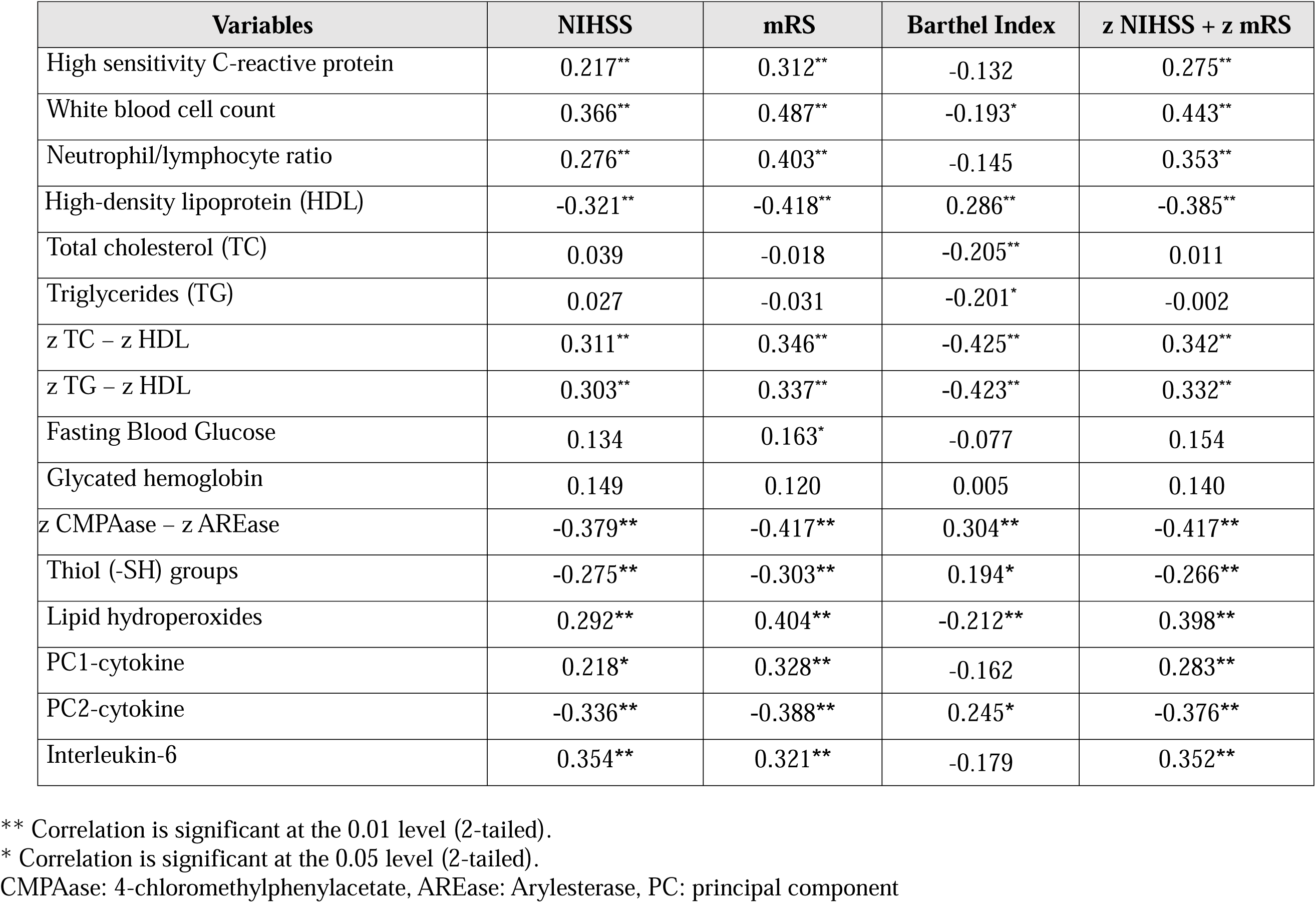
Intercorrelation matrix (Pearson’s correlation coefficients) between biomarkers and severity scales.

The Barthel Index showed a positive correlation with HDL, PC2, z CMPAase – z AREase, and -SH groups. In contrast, significant negative correlations were observed with WBC count, TC, TG, TC/HDL and TG/HDL ratios, and lipid hydroperoxides. The composite z NIHSS + z mRS showed positive correlations with hsCRP, WBC count, NLR, TC/HDL, TG/HDL, lipid hydroperoxides, PC1, and IL-6, whereas negative significant correlations were found with HDL, PC2 score, z CMPAase -z AREase, and -SH groups.

### Diagnostic Performance

We conducted binary logistic regression analyses to identify the strongest predictors of the post-stroke condition, using healthy controls as the reference group and post-stroke patients as the outcome variable. **Table 5** shows two models. Model #1 entered only the MULTIPLEX data and revealed significant associations with PC1 and IL-6 (both positively) and PC2 score and FGF (both inversely). The model produced an effect size of 0.693, resulting in an overall classification accuracy of 87.8%, alongside a sensitivity of 93.4% and a specificity of 71.4%. Model #2, shows that the TG/HDL ratio, z CMPAase -z AREase, and lipid hydroperoxides exhibited significant predictive capability. The model achieved an effect size of 0.803, an overall accuracy of 87.8%, and demonstrated enhanced sensitivity at 95.1% and specificity at 90.5%.

**Table 5:** Results of binary logistic regression analysis with post-stroke patients as dependent variable and healthy controls as reference group.

| Dichotomies |  | Explanatory Variables | B | SE | Wald (df=1) | p | OR | 95% CI |
| --- | --- | --- | --- | --- | --- | --- | --- | --- |
| **Post stroke**<br>**versus healthy**<br>**controls** | **Model #1** | PC1-cytokine | 1.131 | 0.419 | 7.29 | 0.007 | 3.10 | 1.36;7.04 |
|  |  | PC2-cytokine | -2.374 | 0.684 | 12.05 | <0.001 | 0.09 | 0.02;0.36 |
|  |  | Interleukin-6 | 1.297 | 0.534 | 5.91 | 0.015 | 3.66 | 1.29;10.41 |
|  |  | FGF-basic | -0.869 | 0.417 | 4.34 | 0.037 | 0.42 | 0.19;0.95 |
|  | **Model #2** | Atherogenic index of plasma | 1.589 | 0.726 | 4.79 | 0.029 | 4.90 | 1.18;20.34 |
|  |  | z CMPAase – z AREase | -1.861 | 0.568 | 10.74 | 0.001 | 0.16 | 0.05;0.47 |
|  |  | Lipid hydroperoxides | 2.616 | 0.806 | 10.54 | 0.001 | 13.68 | 2.82;66.37 |

CMPAase: 4-chloromethylphenylacetate, AREase: Arylesterase, PC: principal component, FGF: Fibroblast growth factor,

### Prediction of post-stroke severity by NIMETOX biomarkers

**Table 6** displays the results of multivariate regression analyses evaluating the best NIMETOX biomarkers that predict post-stroke severity and functional disability. The dependent variables included NIHSS-2, mRS-2, and the Barthel Index. The explanatory variables comprised inflammatory markers (TNF-α, IL-6, IL-13, PC2-cytokines, NLR, and hsCRP), oxidative stress indices (CMPAase-resPON1 and LOOH1), and metabolic parameters (TC/HDL and HbA1c). In Regression #1, IL-6, HbA1c, and TC/HDL exhibited positive associations, whilst PC2 showed an inverse association and all variables accounted for 44.6% of the variance in the NIHSS score. **Figure 1** presents the partial regression plot, demonstrating the adjusted linear relationship between NIHSS and IL-6. Regression analysis #2 indicated that 55.9% of the variance in mRS was accounted for lipid hydroperoxides, NLR, HbA1c, and hsCRP, all of which exhibited positive associations, alongside z CMPAase – z AREase, which demonstrated a negative association. Since no Multiplex data could be included in this model, we constructed a second model whereby only Multiplex data were entered. IL-6 and PC1 were identified as predictors of mRS, whereas PC2 exhibited an inverse relationship, collectively accounting for 42.2% of the variance. **Figure 2** illustrates the partial regression plot of mRS on IL-6.

**Figure 1.**
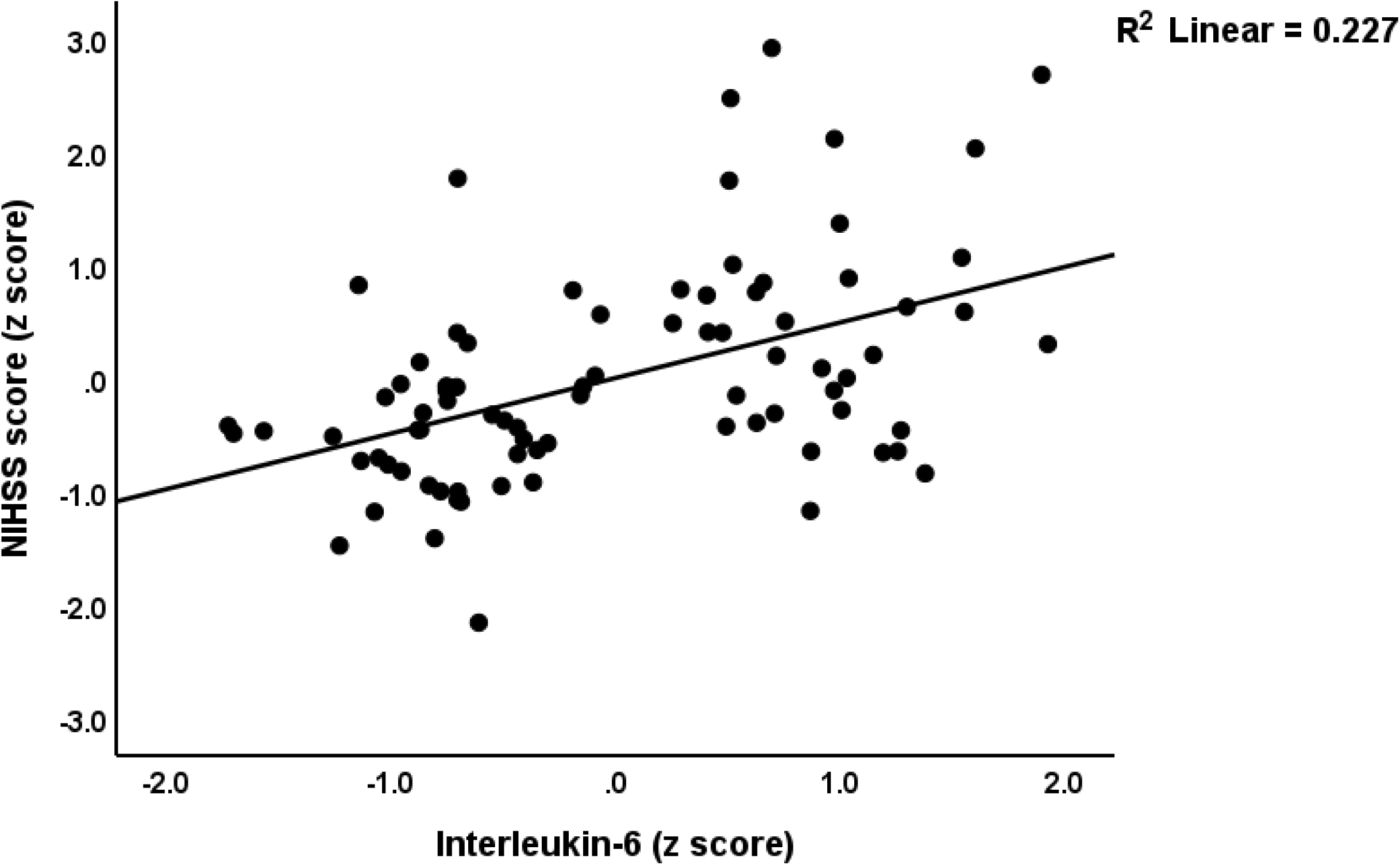
Partial regression plot of the NIH Stroke Scale (NIHSS) score on serum interleukin-6 (p<0.001)

**Figure 2.**
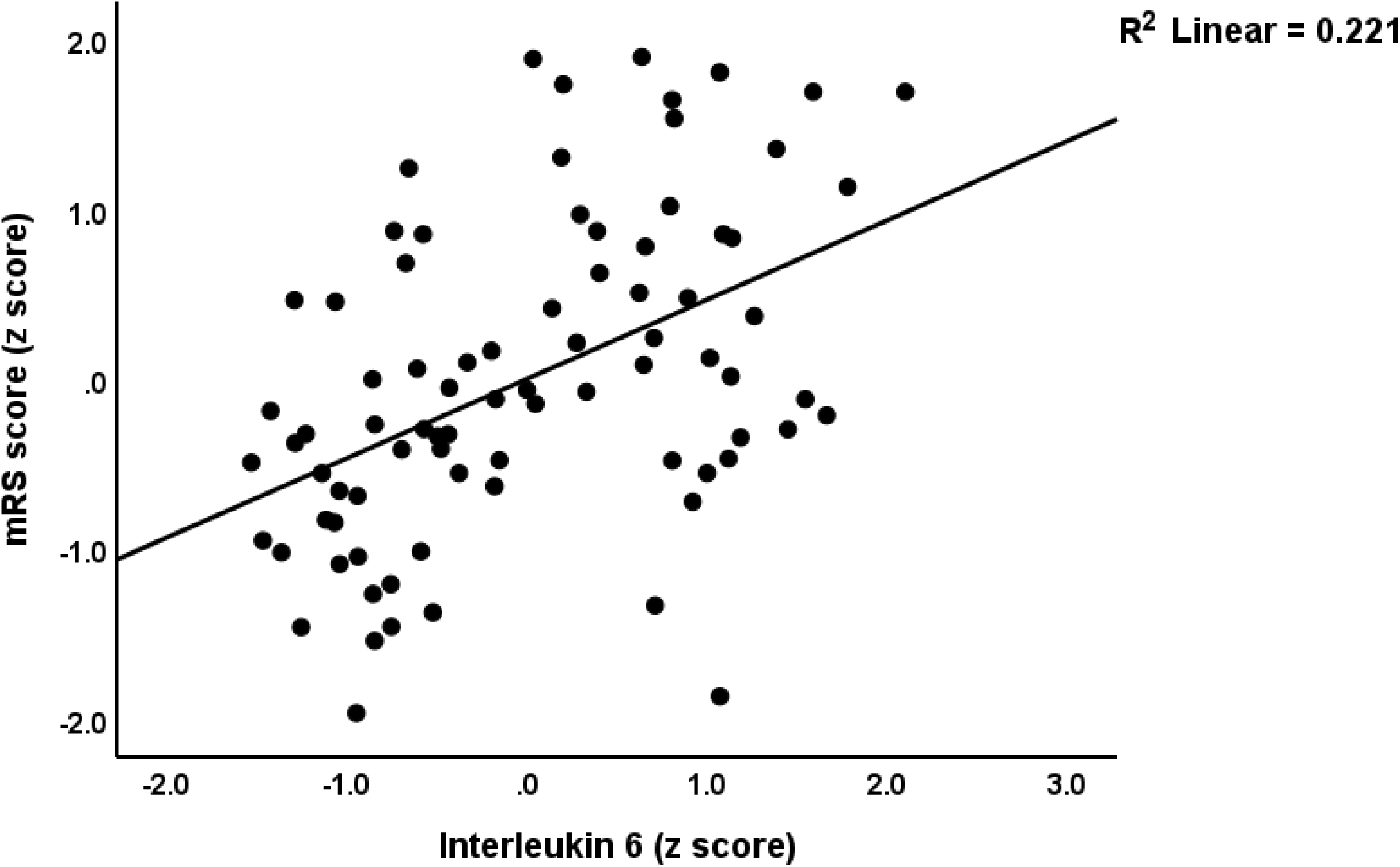
Partial regression plot of the modified Rankin Scale (mRS) on serum interleukin-6 (p<0.001).

**Table 6.** Results of multiple regression analysis with the severity rating scales as dependent variables and biomarkers as explanatory variables.

| Dependent Variables | Explanatory Variables | Coefficient statistics |  |  | Model statistics |  |  |  |
| --- | --- | --- | --- | --- | --- | --- | --- | --- |
| | | $\beta$ | t | p | R<sup>2</sup> | F | df | p |
| #1. NIHSS (all markers are entered) | **Model** |  |  |  | 0.446 | 15.50 | 4/77 | <0.001 |
|  | Castelli risk index 1 | 0.190 | 2.03 | 0.046 |  |  |  |  |
|  | Interleukin-6 | 0.433 | 4.76 | <0.001 |  |  |  |  |
|  | PC2 | -0.432 | -4.74 | <0.001 |  |  |  |  |
|  | HbA1c | 0.228 | 2.54 | 0.013 |  |  |  |  |
| #2. mRS (all biomarkers are entered) | **Model** |  |  |  | 0.559 | 19.29 | 5/76 | <0.001 |
|  | Lipid hydroperoxides | 0.300 | 3.65 | <0.001 |  |  |  |  |
|  | Neutrophil/lymphocyte ratio | 0.339 | 4.34 | <0.001 |  |  |  |  |
|  | z CMPAase – z AREase | -0.325 | -4.05 | <0.001 |  |  |  |  |
|  | HbA1c | 0.218 | 2.85 | 0.006 |  |  |  |  |
|  | hsCRP | 0.198 | 2.51 | 0.014 |  |  |  |  |
| #3. mRS (only Multiplex data are entered) | **Model** |  |  |  | 0.422 | 18.99 | 3/78 | <0.001 |
|  | PC2 | -0.511 | -5.68 | <0.001 |  |  |  |  |
|  | Interleukin-6 | 0.428 | 4.71 | <0.001 |  |  |  |  |
|  | PC1 | 0.263 | 3.02 | 0.003 |  |  |  |  |
| #4. Barthel Index (all variables are entered) | **Model** |  |  |  | 0.531 | 14.16 | 6/75 | <0.001 |
|  | Castelli risk index 1 | -0.366 | -4.46 | <0.001 |  |  |  |  |
|  | z CMPAase – z AREase | 0.199 | 2.09 | 0.040 |  |  |  |  |
|  | Lipid hydroperoxides | -0.213 | -2.50 | 0.015 |  |  |  |  |
| | Tumor necrosis factor-α | -0.302 | -3.48 | <0.001 | | | | |
|  | PC2-cytokine | 0.283 | 2.81 | 0.006 |  |  |  |  |
|  | Interleukin-13 | 0.163 | 2.04 | 0.045 |  |  |  |  |
| #5. Barthel Index (only Multiplex data are entered) | **Model** |  |  |  | 0.285 | 7.67 | 4/77 | <0.001 |
|  | PC2-cytokine | 0.461 | 4.27 | <0.001 |  |  |  |  |
| | Tumor necrosis factor-α | -0.345 | -3.29 | 0.002 | | | | |
|  | Interleukin-6 | -0.292 | -2.85 | 0.006 |  |  |  |  |
|  | Interleukin-13 | 0.238 | 2.44 | 0.017 |  |  |  |  |
| #6. NIHSS (in patients only) | **Model** |  |  |  | 0.272 | 7.11 | 3/57 | <0.001 |
|  | Interleukin-6 | 0.518 | 4.01 | <0.001 |  |  |  |  |
|  | HbA1c | 0.305 | 2.66 | 0.010 |  |  |  |  |
|  | PC2-cytokine | -0.293 | -2.25 | 0.029 |  |  |  |  |
| #7. mRS (in patients only) | **Model** |  |  |  | 0.244 | 9.37 | 2/58 | <0.001 |
|  | Interleukin-6 | 0.347 | 3.02 | 0.004 |  |  |  |  |
|  | Neutrophil/lymphocyte ratio | 0.320 | 2.79 | 0.007 |  |  |  |  |
| #8. Barthel Index (in patients only) | **Model** |  |  |  | 0.184 | 6.52 | 2/58 | 0.003 |
| | Tumor necrosis factor-α | -0.334 | -2.78 | 0.007 | | | | |
|  | Castelli risk index-1 | -0.325 | -2.71 | 0.009 |  |  |  |  |

CMPAase: 4-chloromethylphenylacetate, AREase: Arylesterase, PC: principal component, FGF: Fibroblast growth factor, HbA1c: glycated hemoglobin, hsCRP: high-sensitivity C-reactive protein, NIHSS: National Institutes of Health stroke scale, mRS: Modified Rankin Scale.

Regression #4 model accounted for 53.1% of the variance in the Barthel Index. Positive predictors identified were z CMPAase – z AREase, PC2, and IL-13, whereas z TC – z HDL, lipid hydroperoxides, and TNF-α exhibited inverse associations. Regression #5 entered the Multiplex data only and showed that 28.5% of the variance in the Barthel Index was accounted for by significant positive associations with PC2 and IL-13, alongside inverse associations with IL-6 and TNF-α. **Figure 3** illustrates the partial regression plot of the Barthel index on serum IL-6 levels.

**Figure 3.**
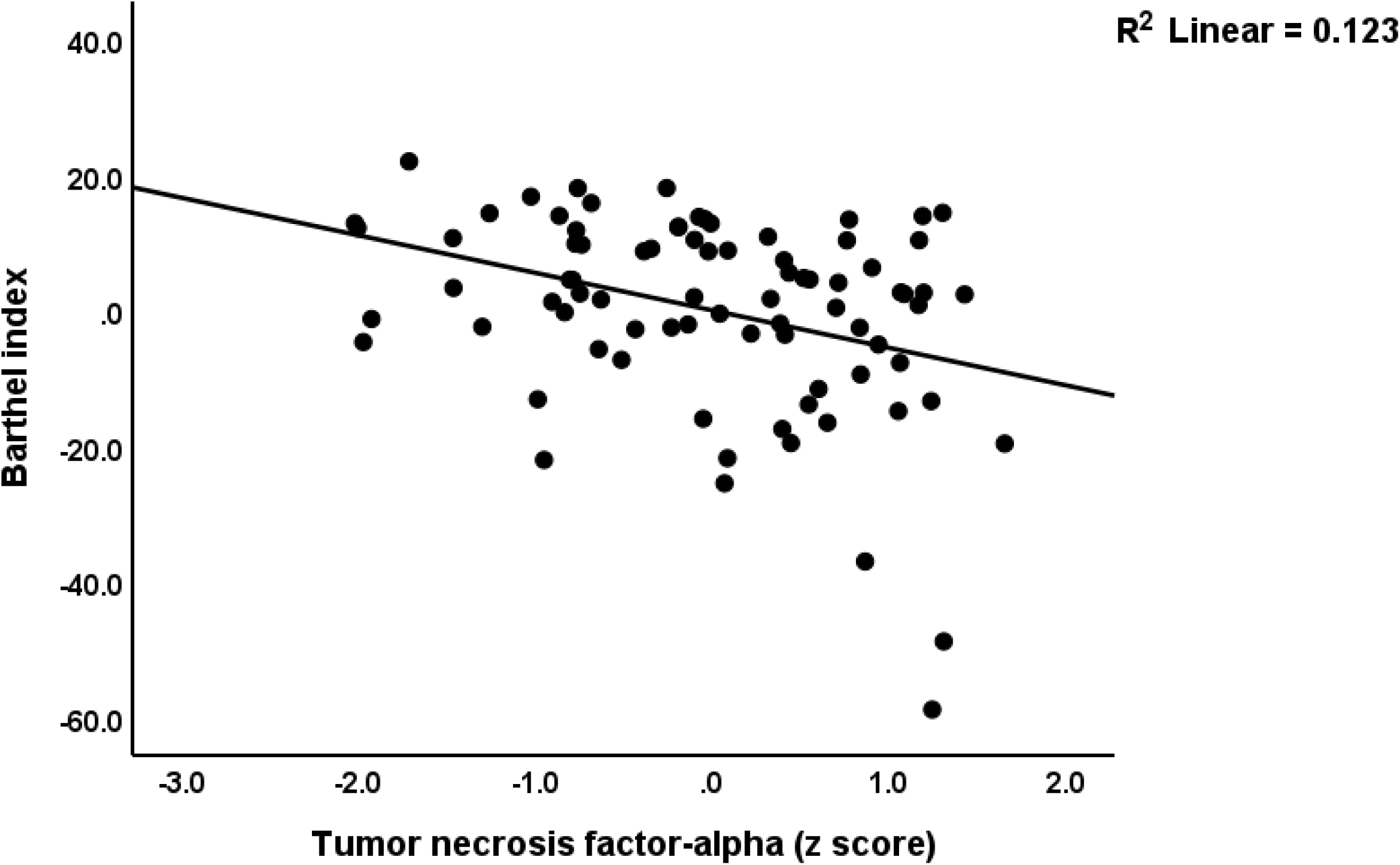
Partial regression plot of the Barthel index on serum tumor necrosis factor-alpha (p<0.001)

Regression models (#6–8) were conducted solely within the post-stroke patient subgroup (controls were excluded). Regression #6 showed that 27.2% of the variance in the NIHSS score was explained by IL-6 and HbA1c (both positively associated) and PC2 (inversely associated). Regression #7 revealed that 24.4% of the variance in the mRS score could be attributed to the effects of IL-6 and NLR, both showing positive associations. Regression #8 demonstrated that 18.4% of the variance in the Barthel Index was predicted by TNF-α and z TC - z HDL, both inversely associated.

Subsequent analyses detailed in Table 2 of the ESF examined predictors of the composite severity scores z NIHSS + z mRS and z NIHSS + z mRS – z Barthel index. In Regression #1, 54.6% of the variance in z NIHSS + z mRS was accounted for by positive associations with NLR, z TC - z HDL, lipid hydroperoxides, and IL-6, alongside inverse associations with z CMPAase – z AREase and PC2. Regression analysis #2 revealed that 59.3% of the variance in the z NIHSS + z mRS – z Barthel composite score was accounted for by the regression on IL-6, TNF-α, z TC – z HDL, and lipid hydroperoxides (all positively correlated), as well as PC2 and z CMPAase - z AREase (both negatively correlated). Regressions #3 and #4 (both performed on the restricted post-stroke sample) indicated that IL-6 alone explained 16.4% of the variance in z NIHSS + z mRS and 14.9% of the variance in z NIHSS + z mRS – z Barthel index, highlighting its reliable predictive significance across various disability measures.

## Discussion

### Abnormal NIMETOX pathways in the post-stroke condition

The current study’s first key finding reveals significant abnormalities in NIMETOX pathways including cytokine networks in post-stroke patients. Disturbances encompass imbalances in proinflammatory cytokines, hematological markers, lipid profiles, glucose metabolism, and oxidative stress indicators, including lipid hydroperoxides, altered nitric oxide metabolism, reduced plasma thiol groups (-SH), and variations in PON1 enzyme activity measured by CMPAase and AREase activities. The results align with previous research identifying immune-inflammatory, metabolic, and oxidative abnormalities after ischemic stroke (Alfieri *et al*., 2020; Brinholi *et al*., 2023; Maes *et al*., 2023). Unlike prior studies that analyzed these elements in isolation, our research provides a comprehensive assessment of these interconnected pathways.

Disruptions in NIMETOX pathways have considerable clinical implications. Sustained inflammatory responses result in heightened neuronal injury, disruption of the blood-brain barrier, and subsequent brain damage (Panickar, 2015; Koyama & Shichita, 2023). Elevated cytokine levels, such as IL-6, IL-1β, and TNF-α, are associated with mood disturbances, cognitive impairment, and fatigue in stroke survivors (Gyawali *et al*., 2020; Ma *et al*., 2024; Zhou *et al*., 2025). However, our study revealed no significant differences in solitary levels of cytokines between post-stroke patients and healthy controls. This finding contrasts with earlier studies linking these cytokines to several neuropsychiatric symptoms, as previously mentioned, and suggests that temporal variability in inflammatory responses may play a role.

The present post-stroke cohort demonstrated notable elevations in fasting blood glucose and HbA1c levels, consistent with the recognized contribution of hyperglycemia to oxidative stress, endothelial dysfunction, and the development of advanced glycation end-products, which further aggravate neurovascular injury (Tomlinson & Gardiner, 2008). Our findings on oxidative stress underscore the biological burden faced by post-stroke patients. LOOH, a notable marker elevated in this cohort, is linked to worse stroke outcomes and insufficient recovery (Maes *et al*., 2023). Mueangson et al. showed that elevated malondialdehyde (another marker of oxidative damage to the lipids) is associated with muscle weakness, and muscle wasting post-stroke (Mueangson *et al*., 2020). Reduced plasma -SH groups suggest a diminished redox buffering capacity, potentially heightening neuronal vulnerability to oxidative damage (Musumeci *et al*., 2013). Diminished nitric oxide bioavailability likely exacerbates this issue by promoting mitochondrial dysfunction and inflammatory signaling (NEMATBAKHSH *et al*., 2008; Ciancarelli *et al*., 2012).

The PON1 enzyme’s functions, indicated by AREase and CMPAase activities, are crucial for antioxidant defense and the regulation of lipid metabolism. The current data indicated an imbalance between the two catalytic sites, with AREase activity surpassing that of CMPAase, which exhibited relatively lower activity. Prior research has demonstrated inverse correlations between AREase levels and stroke risk (Shenhar-Tsarfaty *et al*., 2013; Castellazzi *et al*., 2016), as well as a decrease in CMPAase activity in several neurodegenerative and neuroinflammatory conditions (Moreira *et al*., 2019; Matsumoto *et al*., 2021). The composite score z CMPAase + z HDLc, which indicates the antioxidant and anti-inflammatory properties of the PON1-HDL complex (Maes *et al*., 2018a; Maes *et al*., 2018b), was significantly lower in post-stroke patients. This indicates that compromised PON1-HDL functions might reduce vascular protective mechanisms, consequently enhancing oxidative stress and atherosclerosis (Kotur-Stevuljevic *et al*., 2015; Marsillach *et al*., 2021). In addition to immune and oxidative abnormalities, our patients demonstrate disturbances in lipid profiles, specifically low HDL-C levels alongside increased TC/HDL and TG/HDL ratios. The findings align with previous research linking dyslipidemia to heightened cardiovascular risk, recurrent stroke, and post-stroke depression (Vitturi & Gagliardi, 2022).

The findings indicate that disruptions in NIMETOX pathways significantly contribute to post-stroke pathophysiology, potentially predisposing patients to chronic complications that affect recovery and overall prognosis.

### Prediction of post-stroke patients by NIMETOX components

Our study’s second key finding shows that NIMETOX biomarkers can effectively differentiate post-stroke patients from healthy controls. Two distinct biomarker models were evaluated: one centered on inflammatory cytokines, including PC1- and PC2-cytokines, IL-6, and fibroblast growth factor, and another based on metabolic and oxidative stress biomarkers, specifically the TG/HDL, CMPAase/AREase ratio, and lipid hydroperoxides. The cytokine-focused model attained an overall accuracy of 87.8%, whereas the metabolic and oxidative biomarker model exhibited a higher diagnostic accuracy of 94%. Maes et al. (2023) previously indicated that oxidative stress markers and antioxidant defenses offer enhanced diagnostic potentials relative to immune-inflammatory and metabolic biomarkers, as well as traditional risk factors and MRI stroke volumes in AIS (Maes *et al*., 2023).

Prior research supports our findings by identifying various oxidative and inflammatory biomarkers as significant discriminators of stroke and its complications. Some studies demonstrated high accuracy in classifying IS patients using oxidative stress markers, inflammatory cytokines, glucose, and NOx, in conjunction with traditional risk factors, including sex and blood pressure (Reiche *et al*., 2019; Alfieri *et al*., 2020). In addition, biomarkers such as MDA, IL-6, TNF-α, and intestinal fatty acid binding protein (iFABP), alongside elevated levels of IL-1, IL-4, IL-6, IL-17, IL-18, and decreased IL-10, are effective predictors of post-stroke depression (PSD) (Liu *et al*., 2017; Wang *et al*., 2023; Zheng *et al*., 2024). Yang et al. (2025) identified age, diabetes, hypertension, hs-CRP, and homocysteine as the primary predictors of cognitive impairment post-stroke. A higher granulocyte-to-lymphocyte ratio was recently detected as an independent predictor of early cognitive deficits following a stroke (Huang *et al*., 2025). Kim et al. (2022) identified homocysteine, CRP, total cholesterol, and LDL cholesterol as potential biomarkers for cognitive deficits post-stroke (Kim *et al*., 2022).

Recent studies on biomarkers strengthen the biomarker-diagnostic potential identified in this study. Aldous et al. (2022) utilized a panel of miRNAs, resulting in a diagnostic accuracy of AUC=0.97 for differentiating stroke patients from controls (Aldous *et al*., 2022). Similarly, circRNAs and long non-coding RNAs (lncRNAs) exhibited high accuracy, thereby underscoring the diagnostic importance of molecular biomarkers (Deng *et al*., 2018; Zuo *et al*., 2020). A meta-analysis demonstrated that lncRNAs, especially H19, show significant diagnostic performance for ischemic stroke (AUC=0.88), indicating that molecular biomarkers offer considerable diagnostic value (Pan *et al*., 2024). In addition to these molecular approaches, proteomic studies have further enhanced stroke diagnosis. Misra et al. (2025) identified protein panels that effectively differentiate between stroke subtypes and distinguish true strokes from stroke mimics, with a particular emphasis on proteins involved in inflammation and coagulation pathways (Misra *et al*., 2025). Biomarkers including GFAP, MMP-9, and D-dimer exhibit notable diagnostic capability in differentiating ischemic from hemorrhagic strokes (Kumar *et al*., 2020; Misra *et al*., 2020).

In summary, our study emphasizes the critical diagnostic and prognostic value of a comprehensive assessment of biomarkers in NIMETOX pathways among stroke patients. This integrated approach enhances diagnostic accuracy and provides deeper insights into the biological processes of stroke, facilitating more precise and personalized patient management strategies.

### Association between NIMETOX pathways and post-stroke functional outcomes

The current study’s third key finding indicates that biomarkers in the NIMETOX pathways are significant predictors of stroke severity, disability, and functional outcomes in post-stroke patients. Multiple regression models were employed (either in all participants or in the restricted post-stroke sample) to investigate these associations, each analyzing different combinations of NIMETOX biomarkers.

Stroke severity, as measured by NIHSS scores, was primarily predicted by metabolic markers, including elevated HbA1c and Castelli risk index-1, as well as inflammatory indices such as IL-6 and PC2-cytokine score. The results align with previous studies demonstrating that elevated levels of IL-6 and TNF-α serve as reliable indicators of more severe strokes and unfavorable outcomes (Braadt *et al*., 2023; Băcilă *et al*., 2025). Previous studies have identified lipoprotein(a), ferritin, and AIP, as well as Castelli risk index-1 and 2, as predictors of stroke severity, indicating dyslipidemia (Chakraborty *et al*., 2013; Wang *et al*., 2024). The relationship between elevated HbA1c and stroke severity is supported by evidence indicating higher NIHSS scores in patients with elevated HbA1c (Han *et al*., 2022; Alhawiti *et al*., 2025). Rashid et al. demonstrated a correlation between low NOx levels and heightened stroke severity (Rashid *et al*., 2003).

The present results demonstrate that the severity of post-stroke disabilities can be effectively predicted by various NIMETOX biomarkers, including lipid hydroperoxides, NLR, CMPAase/AREase ratio, HbA1c, and hsCRP. A model concentrating exclusively on immune-inflammatory biomarkers (IL-6, PC1- and PC2-cytokines) explained a substantial portion of the variance in disability outcomes, underscoring the vital role of cytokine networks and immune disbalances in post-stroke disabilities. The results support earlier research indicating that hsCRP and IL-6 serve as independent predictors of disability and negative outcomes following a stroke (Basic Kes *et al*., 2008; Gu *et al*., 2023; M’Barek *et al*., 2025). Meta-analytic evidence indicates that elevated IL-6 levels correlate with an increased mortality risk within three months post-stroke (Zonoozi *et al*., 2021).

The correlation between disturbed PON1 activity, reflected by abnormal CMPAase/AREase ratio, and elevated disability scores aligns with earlier research indicating that impaired CMPAase and AREase activities serve as significant predictors of disability at various intervals following a stroke (Brinholi *et al*., 2023; Maes *et al*., 2023). The predictive significance of LOOH, increased NLR, and dyslipidemia corresponds with previous research associating these markers with post-stroke disability and mortality (Maes *et al*., 2023; Tsalta-Mladenov & Andonova, 2024; Wang *et al*., 2024).

Functional outcomes, assessed with the Barthel index, were associated with a range of NIMETOX biomarkers, including LOOH, the Castelli risk index-1, CMPAase activity, and cytokine networks including TNF-α, IL-13, and PC2-cytokines. Models focusing exclusively on immune biomarkers, including PC2-cytokines, TNF-α, IL-6, and IL-13, supported these associations and explained a substantial portion of the variance in functional recovery. Prior research supports these findings, demonstrating that elevated levels of TNF-α and IL-6 are reliable indicators of diminished functional recovery post-stroke (Li *et al*., 2022; Băcilă *et al*., 2025). Furthermore, IL-6 is correlated with reduced Barthel index scores and increased infarct volumes (Waje-Andreassen *et al*., 2005; Basic Kes *et al*., 2008).

Emerging biomarkers, including neurofilament light (NfL) and glial fibrillary acidic protein (GFAP), have been shown to independently predict functional impairment and negative recovery outcomes following a stroke (Ferrari *et al*., 2023). Elevated serum levels of NfL and GFAP are consistently associated with greater neurological damage and worse clinical outcomes (Uphaus *et al*., 2019; Vollmuth *et al*., 2024), underscoring their prognostic importance. Besides, in patients with moderate to severe NIHSS scores, caspase-3 could be a prognostic indicator of early death (Zaharia *et al*., 2024). Yu H et al. found that the serum level of xanthine oxidase on admission is an independent risk factor for acute ischemic stroke (Yu *et al*., 2023).

Our results highlight the effectiveness of specific NIMETOX pathway biomarkers in elucidating severity, disability, and functional outcomes. The findings provide significant insights into the intricate biological mechanisms involved in stroke recovery and indicate possible directions for enhancing prognostic assessment and individualized post-stroke treatment.

## Limitation

There are certain limitations to the results of this study that should be considered when interpreting them. Adding more NIMETOX biomarkers would enhance the current study’s comprehensiveness. Malondialdehyde, advanced oxidation protein products (AOPP), 8-IsoF2, 4-hydroxynonenal (4-HNE), and tryptophan metabolites are examples of biomarkers. This cross-sectional design study limits our ability to establish a causal association between NIMETOX pathways and post-stroke complications. Thus, future longitudinal studies are required to find the temporal relationships between abnormalities in NIMETOX pathways and stroke outcomes.

## Conclusion

This study indicates that post-stroke conditions are associated with abnormal NIMETOX pathways. The components of these pathways demonstrate diagnostic potential and predictive value regarding stroke severity, disability, and functional outcomes in post-stroke patients. This presents a promising opportunity to enhance the prediction of clinical outcomes and to tailor therapeutic interventions aimed at reducing post-stroke complications and improving recovery.

## Data Availability

The corresponding author (MM) will provide access to the dataset supporting this study upon receipt of a valid request and the completion of a thorough data review.

## Acknowledgements

Not Applicable

## Ethical approval and consent to participate

All participants gave informed consent, and the study was approved by the Institutional Review Board of the Faculty of Medicine, Chulalongkorn University, in accordance with the Declaration of Helsinki and ICH-GCP standards.

## Author’s contributions

MM, TS, and AFA conducted the design of the current study. Samples were collected by TS and CT. The statistical evaluation was conducted by MM. AFA and YZ composed the initial draft which was edited by MM and EB. CT, TS, FB, APM, AKM and DB performed blood analysis. All authors participated in the editing process and have provided their consent for the submission of the final version.

## References

Aldous, E.K., Toor, S.M., Parray, A., Al-Sarraj, Y., Diboun, I., Abdelalim, E.M., Arredouani, A., El-Agnaf, O., Thornalley, P.J., Akhtar, N., Pananchikkal, S.V., Shuaib, A., Alajez, N.M. & Albagha, O.M.E. (2022) Identification of Novel Circulating miRNAs in Patients with Acute Ischemic Stroke. Int J Mol Sci, 23.

Alfieri, D.F., Lehmann, M.F., Flauzino, T., de Araújo, M.C.M., Pivoto, N., Tirolla, R.M., Simão, A.N.C., Maes, M. & Reiche, E.M.V. (2020) Immune-Inflammatory, Metabolic, Oxidative, and Nitrosative Stress Biomarkers Predict Acute Ischemic Stroke and Short-Term Outcome. Neurotoxicity Research, 38, 330–343.

Alghamdi, I., Ariti, C., Williams, A., Wood, E. & Hewitt, J. (2021) Prevalence of fatigue after stroke: A systematic review and meta-analysis. Eur Stroke J, 6, 319–332.

Alhawiti, N.M., Elsokkary, E.M., Aldali, J.A. & Alotaibi, B.A. (2025) Investigating the impact of glycated hemoglobin levels on stroke severity in patients with acute ischemic stroke. Scientific Reports, 15, 12114.

Ali, A., Obaid, O., Akhtar, N., Rao, R., Tora, S.H. & Shuaib, A. (2024) Association between HDL levels and stroke outcomes in the Arab population. Scientific Reports, 14, 3071.

Băcilă, C.I., Vlădoiu, M.G., Văleanu, M., Moga, D.F. & Pumnea, P.M. (2025) The Role of IL-6 and TNF-Alpha Biomarkers in Predicting Disability Outcomes in Acute Ischemic Stroke Patients. Life (Basel), 15.

Basic Kes, V., Simundic, A.M., Nikolac, N., Topic, E. & Demarin, V. (2008) Pro-inflammatory and anti-inflammatory cytokines in acute ischemic stroke and their relation to early neurological deficit and stroke outcome. Clin Biochem, 41, 1330–1334.

Benjamin, E.J., Virani, S.S., Callaway, C.W., Chamberlain, A.M., Chang, A.R., Cheng, S., Chiuve, S.E., Cushman, M., Delling, F.N., Deo, R., de Ferranti, S.D., Ferguson, J.F., Fornage, M., Gillespie, C., Isasi, C.R., Jiménez, M.C., Jordan, L.C., Judd, S.E., Lackland, D., Lichtman, J.H., Lisabeth, L., Liu, S., Longenecker, C.T., Lutsey, P.L., Mackey, J.S., Matchar, D.B., Matsushita, K., Mussolino, M.E., Nasir, K., O’Flaherty, M., Palaniappan, L.P., Pandey, A., Pandey, D.K., Reeves, M.J., Ritchey, M.D., Rodriguez, C.J., Roth, G.A., Rosamond, W.D., Sampson, U.K.A., Satou, G.M., Shah, S.H., Spartano, N.L., Tirschwell, D.L., Tsao, C.W., Voeks, J.H., Willey, J.Z., Wilkins, J.T., Wu, J.H., Alger, H.M., Wong, S.S. & Muntner, P. (2018) Heart Disease and Stroke Statistics-2018 Update: A Report From the American Heart Association. Circulation, 137, e67–e492.

Braadt, L., Naumann, M., Freuer, D., Schmitz, T., Linseisen, J. & Ertl, M. (2023) Novel inflammatory biomarkers associated with stroke severity: results from a cross-sectional stroke cohort study. Neurological Research and Practice, 5, 31.

Brinholi, F.F., Michelin, A.P., Matsumoto, A.K., de O Semeão, L., Almulla, A.F., Supasitthumrong, T., Tunvirachaisakul, C., Barbosa, D.S. & Maes, M. (2023) Paraoxonase 1 status is a major Janus-faced component of mild and moderate acute ischemic stroke and consequent disabilities. Metabolic Brain Disease, 38, 2115–2131.

Castellazzi, M., Trentini, A., Romani, A., Valacchi, G., Bellini, T., Bonaccorsi, G., Fainardi, E., Cavicchio, C., Passaro, A., Zuliani, G. & Cervellati, C. (2016) Decreased arylesterase activity of paraoxonase-1 (PON-1) might be a common denominator of neuroinflammatory and neurodegenerative diseases. Int J Biochem Cell Biol, 81, 356–363.

Chakraborty, B., Vishnoi, G., Goswami, B., Gowda, S.H., Chowdhury, D. & Agarwal, S. (2013) Lipoprotein(a), ferritin, and albumin in acute phase reaction predicts severity and mortality of acute ischemic stroke in North Indian Patients. J Stroke Cerebrovasc Dis, 22, e159–167.

Ciancarelli, I., De Amicis, D., Di Massimo, C., Carolei, A. & Ciancarelli, M.G. (2012) Oxidative stress in post-acute ischemic stroke patients after intensive neurorehabilitation. Curr Neurovasc Res, 9, 266–273.

Deng, Q.W., Li, S., Wang, H., Sun, H.L., Zuo, L., Gu, Z.T., Lu, G., Sun, C.Z., Zhang, H.Q. & Yan, F.L. (2018) Differential long noncoding RNA expressions in peripheral blood mononuclear cells for detection of acute ischemic stroke. Clin Sci (Lond), 132, 1597–1614.

Feigin, V.L., Krishnamurthi, R.V., Parmar, P., Norrving, B., Mensah, G.A., Bennett, D.A., Barker-Collo, S., Moran, A.E., Sacco, R.L., Truelsen, T., Davis, S., Pandian, J.D., Naghavi, M., Forouzanfar, M.H., Nguyen, G., Johnson, C.O., Vos, T., Meretoja, A., Murray, C.J.L., Roth, G.A., Group, G.W. & Group, G.S.P.E. (2015) Update on the Global Burden of Ischemic and Hemorrhagic Stroke in 1990-2013: The GBD 2013 Study. Neuroepidemiology, 45, 161–176.

Ferrari, F., Rossi, D., Ricciardi, A., Morasso, C., Brambilla, L., Albasini, S., Vanna, R., Fassio, C., Begenisic, T., Loi, M., Bossi, D., Zaliani, A., Alberici, E., Lisi, C., Morotti, A., Cavallini, A., Mazzacane, F., Nardone, A., Corsi, F. & Truffi, M. (2023) Quantification and prospective evaluation of serum NfL and GFAP as blood-derived biomarkers of outcome in acute ischemic stroke patients. J Cereb Blood Flow Metab, 43, 1601–1611.

Ferretti, G., Bacchetti, T., Masciangelo, S., Nanetti, L., Mazzanti, L., Silvestrini, M., Bartolini, M. & Provinciali, L. (2008) Lipid peroxidation in stroke patients. Clinical Chemistry and Laboratory Medicine, 46, 113–117.

Furlan, J.C., Vergouwen, M.D.I., Fang, J. & Silver, F.L. (2014) White blood cell count is an independent predictor of outcomes after acute ischaemic stroke. European Journal of Neurology, 21, 215–222.

Gu, H.Q., Yang, K.X., Li, J.J., Lin, J.X., Jing, J., Xiong, Y.Y., Zhao, X.Q., Wang, Y.L., Liu, L.P., Meng, X., Jiang, Y., Li, H., Wang, Y.J. & Li, Z.X. (2023) Mediation effect of stroke recurrence in the association between post-stroke interleukin-6 and functional disability. CNS Neurosci Ther, 29, 3579–3587.

Gyawali, P., Hinwood, M., Chow, W.Z., Kluge, M., Ong, L.K., Nilsson, M. & Walker, F.R. (2020) Exploring the relationship between fatigue and circulating levels of the pro-inflammatory biomarkers interleukin-6 and C-reactive protein in the chronic stage of stroke recovery: A cross-sectional study. Brain, Behavior, & Immunity - Health, 9, 100157.

Han, L., Hou, Z., Ma, M., Ding, D., Wang, D. & Fang, Q. (2022) Impact of glycosylated hemoglobin on early neurological deterioration in acute mild ischemic stroke patients treated with intravenous thrombolysis. Front Aging Neurosci, 14, 1073267.

Hu, M.-L. (1994) [41] Measurement of protein thiol groups and glutathione in plasma Methods in Enzymology. Academic Press, pp. 380–385.

Huang, W., Liao, L., Liu, Q., Ma, R., Hu, W., Dai, Y., Wang, L. & Sha, D. (2025) Predictive value of circulating inflammatory biomarkers for early-onset post-stroke cognitive impairment: a prospective cohort study. Front Neurol, 16, 1565613.

Katan, M. & Luft, A. (2018) Global Burden of Stroke. Semin Neurol, 38, 208–211.

Kawabori, M. & Yenari, M.A. (2015) Inflammatory responses in brain ischemia. Curr Med Chem, 22, 1258–1277.

Kim, K.Y., Shin, K.Y. & Chang, K.-A. (2022) Potential Biomarkers for Post-Stroke Cognitive Impairment: A Systematic Review and Meta-Analysis International Journal of Molecular Sciences.

Koton, S., Tanne, D., Green, M.S. & Bornstein, N.M. (2010) Mortality and predictors of death 1 month and 3 years after first-ever ischemic stroke: data from the first national acute stroke Israeli survey (NASIS 2004). Neuroepidemiology, 34, 90–96.

Kotur-Stevuljevic, J., Bogavac-Stanojevic, N., Jelic-Ivanovic, Z., Stefanovic, A., Gojkovic, T., Joksic, J., Sopic, M., Gulan, B., Janac, J. & Milosevic, S. (2015) Oxidative stress and paraoxonase 1 status in acute ischemic stroke patients. Atherosclerosis, 241, 192–198.

Koyama, R. & Shichita, T. (2023) Glial roles in sterile inflammation after ischemic stroke. Neurosci Res, 187, 67–71.

Kumar, A., Misra, S., Yadav, A.K., Sagar, R., Verma, B., Grover, A. & Prasad, K. (2020) Role of glial fibrillary acidic protein as a biomarker in differentiating intracerebral haemorrhage from ischaemic stroke and stroke mimics: a meta-analysis. Biomarkers, 25, 1–8.

Lehmann, A., Alfieri, D.F., de Araújo, M.C.M., Trevisani, E.R., Nagao, M.R., Pesente, F.S., Gelinski, J.R., de Freitas, L.B., Flauzino, T., Lehmann, M.F., Lozovoy, M.A.B., Breganó, J.W., Simão, A.N.C., Maes, M. & Reiche, E.M.V. (2022) Immune-inflammatory, coagulation, adhesion, and imaging biomarkers combined in machine learning models improve the prediction of death 1 year after ischemic stroke. Clin Exp Med, 22, 111–123.

Li, J., Lin, J., Pan, Y., Wang, M., Meng, X., Li, H., Wang, Y., Zhao, X., Qin, H., Liu, L. & Wang, Y. (2022) Interleukin-6 and YKL-40 predicted recurrent stroke after ischemic stroke or TIA: analysis of 6 inflammation biomarkers in a prospective cohort study. J Neuroinflammation, 19, 131.

Liu, Z., Zhu, Z., Zhao, J., Ren, W., Cai, Y., Wang, Q., Luan, X., Zhao, K. & He, J. (2017) Malondialdehyde: A novel predictive biomarker for post-stroke depression. Journal of Affective Disorders, 220, 95–101.

M’Barek, L., Jin, A., Pan, Y., Lin, J., Jiang, Y., Meng, X. & Wang, Y. (2025) Stroke Prognosis: The Impact of Combined Thrombotic, Lipid, and Inflammatory Markers. J Atheroscler Thromb, 32, 458–473.

Ma, Y., Chen, Y., Yang, T., He, X., Yang, Y., Chen, J. & Han, L. (2024) Blood biomarkers for post-stroke cognitive impairment: A systematic review and meta-analysis. Journal of Stroke and Cerebrovascular Diseases, 33.

Maes, M., Almulla, A.F., You, Z. & Zhang, Y. (2025a) Neuroimmune, metabolic and oxidative stress pathways in major depressive disorder. Nature Reviews Neurology.

Maes, M., Bonifacio, K.L., Morelli, N.R., Vargas, H.O., Moreira, E.G., St. Stoyanov, D., Barbosa, D.S., Carvalho, A.F. & Nunes, S.O.V. (2018a) Generalized anxiety disorder (GAD) and comorbid major depression with GAD are characterized by enhanced nitro-oxidative stress, increased lipid peroxidation, and lowered lipid-associated antioxidant defenses. Neurotoxicity research, 34, 489–510.

Maes, M., Brinholi, F.F., Michelin, A.P., Matsumoto, A.K., de Oliveira Semeão, L., Almulla, A.F., Supasitthumrong, T., Tunvirachaisakul, C. & Barbosa, D.S. (2023) In Mild and Moderate Acute Ischemic Stroke, Increased Lipid Peroxidation and Lowered Antioxidant Defenses Are Strongly Associated with Disabilities and Final Stroke Core Volume. Antioxidants, 12, 188.

Maes, M., Congio, A., Moraes, J.B., Bonifacio, K.L., Barbosa, D.S., Vargas, H.O., Morris, G., Puri, B.K., Michelin, A.P. & Nunes, S.O.V. (2018b) Early life trauma predicts affective phenomenology and the effects are partly mediated by staging coupled with lowered lipid-associated antioxidant defences. Biomolecular concepts, 9, 115–130.

Maes, M., Jirakran, K., Semeão, L.d.O., Michelin, A.P., Matsumoto, A.K., Brinholi, F.F., Barbosa, D.S., Tivirachaisakul, C., Almulla, A.F., Stoyanov, D. & Zhang, Y. (2025b) Key factors underpinning neuroimmune-metabolic-oxidative (NIMETOX) major depression in outpatients: paraoxonase 1 activity, reverse cholesterol transport, increased atherogenicity, protein oxidation, and differently expressed cytokine networks. medRxiv, 2025.2003.2002.25323183.

Maes, M., Jirakran, K., Vasupanrajit, A., Zhou, B., Tunvirachaisakul, C., Stoyanov, D.S. & Almulla, A.F. (2024) Are abnormalities in lipid metabolism, together with adverse childhood experiences, the silent causes of immune-linked neurotoxicity in major depression? Neuro Endocrinol Lett, 45, 393–408.

Manolescu, B.N., Berteanu, M., Oprea, E., Chiriac, N., Dumitru, L., Vlădoiu, S., Popa, O. & Ianăş, O. (2011) Dynamic of oxidative and nitrosative stress markers during the convalescent period of stroke patients undergoing rehabilitation. Annals of Clinical Biochemistry, 48, 338–343.

Marsillach, J., Richter, R.J., Costa, L.G. & Furlong, C.E. (2021) Paraoxonase-1 (PON1) Status Analysis Using Non-Organophosphate Substrates. Curr Protoc, 1, e25.

Matsumoto, A.K., Maes, M., Supasitthumrong, T., Maes, A., Michelin, A.P., de Oliveira Semeão, L., de Lima Pedrão, J.V., Moreira, E.G., Kanchanatawan, B. & Barbosa, D.S. (2021) Deficit schizophrenia and its features are associated with PON1 Q192R genotypes and lowered paraoxonase 1 (PON1) enzymatic activity: effects on bacterial translocation. CNS Spectr, 26, 406–415.

Meschia, J.F. & Brott, T. (2018) Ischaemic stroke. European Journal of Neurology, 25, 35–40.

Misra, S., Montaner, J., Ramiro, L., Arora, R., Talwar, P., Nath, M., Kumar, A., Kumar, P., Pandit, A.K., Mohania, D., Prasad, K. & Vibha, D. (2020) Blood biomarkers for the diagnosis and differentiation of stroke: A systematic review and meta-analysis. Int J Stroke, 15, 704–721.

Misra, S., Natu, A., Kumar, P., Watson, C., Frankel, M. & Rangaraju, S. (2025) Abstract WMP32: Cross-Platform Proteomics and Machine Learning Algorithms Nominate Biomarkers of Stroke Diagnosis: An Exploratory Study. Stroke, 56, AWMP32–AWMP32.

Mitchell, A.J., Sheth, B., Gill, J., Yadegarfar, M., Stubbs, B., Yadegarfar, M. & Meader, N. (2017) Prevalence and predictors of post-stroke mood disorders: A meta-analysis and meta-regression of depression, anxiety and adjustment disorder. Gen Hosp Psychiatry, 47, 48–60.

Moreira, E.G., Correia, D.G., Bonifácio, K.L., de Moraes, J.B., Cavicchioli, F.L., Nunes, C.S., Nunes, S.O.V., Vargas, H.O., Barbosa, D.S. & Maes, M. (2019) Lowered PON1 activities are strongly associated with depression and bipolar disorder, recurrence of (hypo) mania and depression, increased disability and lowered quality of life. The World Journal of Biological Psychiatry.

Mueangson, O., Vongvaivanichakul, P., Kamdee, K., Jansakun, C., Chulrik, W., Pongpanitanont, P., Sathirapanya, P. & Chunglok, W. (2020) Malondialdehyde as a Useful Biomarker of Low Hand Grip Strength in Community-Dwelling Stroke Patients. Int J Environ Res Public Health, 17.

Musumeci, M., Sotgiu, S., Persichilli, S., Arru, G., Angeletti, S., Fois, M.L., Minucci, A. & Musumeci, S. (2013) Role of SH levels and markers of immune response in the stroke. Dis Markers, 35, 141–147.

Nematbakhsh, M., Ashtari, F., Mousavi, M.E., Nouranian, E. & Saadatnia, M. (2008) The correlation of nitrite concentration with lesion size in initial phase of stroke; It is not correlated with National Institute Health Stroke Scale.

Nguyen, V.A., Crewther, S.G., Howells, D.W., Wijeratne, T., Ma, H., Hankey, G.J., Davis, S., Donnan, G.A. & Carey, L.M. (2020) Acute Routine Leukocyte and Neutrophil Counts Are Predictive of Poststroke Recovery at 3 and 12 Months Poststroke: An Exploratory Study. Neurorehabilitation and Neural Repair, 34, 844–855.

Pan, J., Fan, W., Gu, C., Xi, Y., Wang, Y. & Wang, P. (2024) Long Non-Coding RNAs as Diagnostic Biomarkers for Ischemic Stroke: A Systematic Review and Meta-Analysis Genes.

Panickar, K.S. (2015) Chapter 1 - Anti-Inflammatory Properties of Botanical Extracts Contribute to Their Protective Effects in Brain Edema in Cerebral Ischemia. In Watson, R.R., Preedy, V.R. (eds) Bioactive Nutraceuticals and Dietary Supplements in Neurological and Brain Disease. Academic Press, San Diego, pp. 3–15.

Rashid, P.A., Whitehurst, A., Lawson, N. & Bath, P.M.W. (2003) Plasma nitric oxide (nitrate/nitrite) levels in acute stroke and their relationship with severity and outcome. Journal of Stroke and Cerebrovascular Diseases, 12, 82–87.

Reiche, E.M.V., Gelinksi, J.R., Alfieri, D.F., Flauzino, T., Lehmann, M.F., de Araújo, M.C.M., Lozovoy, M.A.B., Simão, A.N.C., de Almeida, E.R.D. & Maes, M. (2019) Immune-inflammatory, oxidative stress and biochemical biomarkers predict short-term acute ischemic stroke death. Metab Brain Dis, 34, 789–804.

Shaafi, S., Sharifipour, E., Rahmanifar, R., Hejazi, S., Andalib, S., Nikanfar, M., Baradarn, B. & Mehdizadeh, R. (2014) Interleukin-6, a reliable prognostic factor for ischemic stroke. Iran J Neurol, 13, 70–76.

Shenhar-Tsarfaty, S., Waiskopf, N., Ofek, K., Shopin, L., Usher, S., Berliner, S., Shapira, I., Bornstein, N.M., Ritov, Y., Soreq, H. & Ben Assayag, E. (2013) Atherosclerosis and arteriosclerosis parameters in stroke patients associate with paraoxonase polymorphism and esterase activities. Eur J Neurol, 20, 891–898.

Simats, A. & Liesz, A. (2022) Systemic inflammation after stroke: implications for post-stroke comorbidities. EMBO Molecular Medicine, 14, e16269.

Smith, C.J., Emsley, H.C., Gavin, C.M., Georgiou, R.F., Vail, A., Barberan, E.M., del Zoppo, G.J., Hallenbeck, J.M., Rothwell, N.J., Hopkins, S.J. & Tyrrell, P.J. (2004) Peak plasma interleukin-6 and other peripheral markers of inflammation in the first week of ischaemic stroke correlate with brain infarct volume, stroke severity and long-term outcome. BMC Neurol, 4, 2.

Taylan, E. & Resmi, H. (2010) The analytical performance of a microplate method for total sulfhydryl measurement in biological samples. TURKISH JOURNAL OF BIOCHEMISTRY-TURK BIYOKIMYA DERGISI, 35, 275–278.

Tomlinson, D.R. & Gardiner, N.J. (2008) Glucose neurotoxicity. Nature Reviews Neuroscience, 9, 36–45.

Tsalta-Mladenov, M.E. & Andonova, S.P. (2024) Peripheral blood cell count ratios as a predictor of poor functional outcome in patients with acute ischemic stroke. Neurol Res, 46, 213–219.

Uphaus, T., Bittner, S., Gröschel, S., Steffen, F., Muthuraman, M., Wasser, K., Weber-Krüger, M., Zipp, F., Wachter, R. & Gröschel, K. (2019) NfL (Neurofilament Light Chain) Levels as a Predictive Marker for Long-Term Outcome After Ischemic Stroke. Stroke, 50, 3077–3084.

Vila, N., Castillo, J., Dávalos, A. & Chamorro, A. (2000) Proinflammatory cytokines and early neurological worsening in ischemic stroke. Stroke, 31, 2325–2329.

Vitturi, B.K. & Gagliardi, R.J. (2022) The prognostic significance of the lipid profile after an ischemic stroke. Neurol Res, 44, 139–145.

Vollmuth, C., Fiessler, C., Montellano, F.A., Kollikowski, A.M., Essig, F., Oeckl, P., Barba, L., Steinacker, P., Schulz, C., Ungethüm, K., Wolf, J., Pham, M., Schuhmann, M.K., Heuschmann, P.U., Haeusler, K.G., Stoll, G., Otto, M. & Neugebauer, H. (2024) Incremental value of serum neurofilament light chain and glial fibrillary acidic protein as blood-based biomarkers for predicting functional outcome in severe acute ischemic stroke. Eur Stroke J, 9, 751–762.

Waje-Andreassen, U., Kråkenes, J., Ulvestad, E., Thomassen, L., Myhr, K.M., Aarseth, J. & Vedeler, C.A. (2005) IL-6: an early marker for outcome in acute ischemic stroke. Acta Neurol Scand, 111, 360–365.

Wang, L., Chunyou, C., Zhu, J., Bao, X. & Tao, X. (2023) Prediction of post-stroke depression with combined blood biomarkers IL-6, TNF-a, and fatty acid binding protein: A prospective study. J Med Biochem, 42, 638–644.

Wang, X., Wu, L., Shu, P., Yu, W. & Yu, W. (2024) Significant association between three atherosclerosis indexes and stroke risk. PLoS One, 19, e0315396.

Weterings, R.P., Kessels, R.P., de Leeuw, F.E. & Piai, V. (2023) Cognitive impairment after a stroke in young adults: A systematic review and meta-analysis. Int J Stroke, 18, 888–897.

Whiteley, W., Jackson, C., Lewis, S., Lowe, G., Rumley, A., Sandercock, P., Wardlaw, J., Dennis, M. & Sudlow, C. (2009) Inflammatory markers and poor outcome after stroke: a prospective cohort study and systematic review of interleukin-6. PLoS Med, 6, e1000145.

Yeh, P.-S., Yang, C.-M., Lin, S.-H., Wang, W.-M., Chen, P.-S., Chao, T.-H., Lin, H.-J., Lin, K.-C., Chang, C.-Y., Cheng, T.-J. & Li, Y.-H. (2013) Low levels of high-density lipoprotein cholesterol in patients with atherosclerotic stroke: A prospective cohort study. Atherosclerosis, 228, 472–477.

Yu, H., Chen, X., Guo, X., Chen, D., Jiang, L., Qi, Y., Shao, J., Tao, L., Hang, J., Lu, G., Chen, Y. & Li, Y. (2023) The clinical value of serum xanthine oxidase levels in patients with acute ischemic stroke. Redox Biol, 60, 102623.

Zaharia, A.-L., Oprea, V.D., Coadă, C.A., Tutunaru, D., Romila, A., Stan, B., Croitoru, A., Ionescu, A.-M. & Lungu, M. (2024) Serum Caspase-3 Levels as a Predictive Molecular Biomarker for Acute Ischemic Stroke. International Journal of Molecular Sciences, 25.

Zheng, T., Jiang, T., Li, R., Zhu, Y., Han, Q. & Wang, M. (2024) Circulating interleukins concentrations and post-stroke depression: A systematic review and meta-analysis. Progress in Neuro-Psychopharmacology and Biological Psychiatry, 134, 111050.

Zhou, Y., Zhao, L., Tang, Y. & Qian, S. (2025) Association between serum inflammatory cytokines levels and post-stroke depression among stroke patients: A meta-analysis and systematic review. Journal of Psychosomatic Research, 190, 112050.

Zonoozi, A.K., Tsarouhas, K., Sotoudeh, M.M., Avan, A., Rezaee, R. & Morovatdar, N. (2021) Interleukin-6 levels predicts mortality after stroke: A systematic review and meta-analysis. Farmacia, 69, 642–649.

Zuo, L., Zhang, L., Zu, J., Wang, Z., Han, B., Chen, B., Cheng, M., Ju, M., Li, M., Shu, G., Yuan, M., Jiang, W., Chen, X., Yan, F., Zhang, Z. & Yao, H. (2020) Circulating Circular RNAs as Biomarkers for the Diagnosis and Prediction of Outcomes in Acute Ischemic Stroke. Stroke, 51, 319–323.

